# Factors influencing modern contraceptive use in Kinshasa, Democratic Republic of Congo

**DOI:** 10.1101/2023.01.31.23285289

**Authors:** Reagan Ingoma Mokeke, Whiejong M. Han, Hee Cheol Kang, Han Nah Kim

## Abstract

Family planning allows individuals and couples to schedule and regulate births at the appropriate time and number. Women who carry unplanned pregnancies are susceptible to postnatal complications, which directly impact their well-being, making them a burden to society. There are 27600 adolescent abortions in the Democratic Republic of the Congo (DRC), accounting for about 19 percent of abortion cases in Kinshasa in 2021 while the reproduction rate remains one of the highest in the world at 5.7 and a low contraceptive prevalence across all methods (28%) in 2022. This study aims to determine the factors that influence modern contraceptive use among women in Kinshasa city in the Democratic Republic of Congo by using data from the PMA 2020 cross-sectional survey to assess the association between predictors and modern contraceptive use through logistic regression analysis (OR and 95% CI).

Findings show that older women are more likely to use modern contraceptive than younger (15-22) and their sensitivity diminishes as they age. This likelihood is also high when women do not discuss this topic with their partner. As household size increases, women’s use of modern contraceptives is positively affected (OR: 4.993). The same is true for women who have given birth at least compared to those who have never given birth (OR: 2.313). Wanting fewer children makes her more likely to use these contraceptives than when she wants more (OR: 1.244). Male condoms, emergency contraceptives and pills were used more by women aged 15-31 while the most common contraceptive method used only by older women is female sterilization.

In a country with rapid population growth such as the DRC, the strategy should focus on raising awareness among young women through school programs, health facilities, community outreach, and other means to achieve optimal use of modern contraceptives based on commonly used methods and involving adult men and women.

## 1. INTRODUCTION

Family planning allows individuals and couples to schedule and regulate births at the appropriate time and number according to the WHO. It is made possible through the application of natural or artificial contraception. Family planning offers the couple and especially the woman a state of overall well-being, with measurable benefits on her reproductive health. (1)

The concept of contraception goes back to long before Jesus Christ in the tribe of Israel(2)

The world’s population grew from about 1.7 billion to 6 billion between 1900 and 1999. The least developed countries account for most of this population and could reach 90% of the world population by 2050.(3)

An estimated 222 million women worldwide are unable to plan a pregnancy despite their intention to do so(4). The most likely causes are those related to the availability of and access to contraception, limited choice of contraceptive method, ethnic, cultural and religious factors, side effects and previous reproductive health experience.

The rate of maternal death and unsafe abortion remains very high in developing countries, with nearly 21 million abortions and 47,000 maternal deaths (5). The practice of family planning could help lower the mortality curve and achieve the Sustainable Development Goals by promoting access to contraception for all to ensure a state of complete well-being and community development as well as women’s empowerment. Successful contraception leads to direct benefits for maternal and child health, including the prevention of unwanted pregnancies and all that goes with it.

Several studies show that in the long term, when a pregnancy is unplanned, it exposes the woman to inappropriate obstetrical management than when it is planned. In addition, there are maternal-fetal complications of varying degrees depending on the age of the pregnancy (6).

Each year, approximately 23 million girls aged 15-19 in poor countries have an unmet need for modern contraception, the majority of whom become pregnant before reaching age 15. Worldwide, 885 million women of childbearing age express a desire for contraception.(7)

In sub-Saharan Africa, contraceptive promotion remains a major challenge. The prevalence of modern contraceptive use has not shown significant variation over time. These variations are 0.77% in Lagos, Nigeria, and 3.64% in Ghana. (8)

The Democratic Republic of the Congo (DRC) is the third-largest country in sub-Saharan Africa, after Algeria and Sudan, with an area of 2,345,000 km^2^ its capital is Kinshasa with a total population of approximately 92,377,986 in 2021 of which 50.1% will be female and 45% under 15 years old. It is estimated that the population will reach 150 million by 2050 with an estimated current GDP per population of 584.1 in 2021 and a life expectancy of 61 years (9). Kinshasa is a megalopolis located in the west of the DRC. Together with Brazzaville, they form the two closest capitals in the world. Its population is estimated at 15,628,085 inhabitants with an average annual growth rate of 4% and a projection of 26 million inhabitants by 2030 (10), In 2010, about 23% of girls (15-19 years) had a live birth in Kinshasa (11).

According to the main indicators of family planning in 2020, in DRC, the unmet need for modern contraception is about 40.2%, the number of unwanted pregnancies is 1,918,000. while the Sustainable Development Goals call for wider health care coverage, including reproductive health. (12)

The DRC has a mix of strong family planning achievements and extreme family planning challenges. and is focusing on introducing new advances such as self-injection for Depot medroxyprogesterone acetate Subcutaneous. A number of provinces are still experiencing armed conflict and others are landlocked, resulting in limited or no access to family planning products and/or services, as well as supply chain issues. In order to advance family planning, the country is focusing on the government’s acceptance and approval of its Reproductive Health and Family Planning Law, which aims to establish and fund a budget line for family planning and increase access to FP services and supplies for all, including youth.

The national strategy 2021-2025 is to provide all Congolese of reproductive age with access to quality, affordable family planning services, regardless of social class, geographic location, or political or religious affiliation (13). Despite government support, family planning still faces major challenges in the DRC, mainly contraceptive security, which consists of maintaining a satisfactory level of flow of contraceptives to the country’s clinical facilities requiring these products, thanks to the selection of commodities, anticipation of needs, supply, delivery, storage, distribution and evaluation of this entire process.

DRC is undoubtedly one of the most multi-ethnic countries in Africa, with people of many origins, cultures and speaking different languages. The use of modern contraceptive methods by women particularly in Kinshasa, is influenced by several factors and at different levels. The law requires the consent of the couple before any implementation of a method of contraception. In case of disagreement between spouses, the position of the spouse concerned by the method to be used takes precedence.(14)

In general, customary, religious and cultural considerations prevail over the idea of restricting pregnancy or maternity. Children are considered a gift from God, a blessing for the couple. Contraceptive methods are sometimes seen as a Western way of controlling local populations. This perception of birth control makes it difficult for family planning to be more effective, especially in rural areas. Some religions, such as Islam, which enshrines polygamy, place women in a situation of competition with each other so that the woman who has more children believes she has more attention or benefits from her husband. In some ethnic groups in the central DRC, having more children is a sign of wealth, because even when one has no means to support oneself, one hopes that having more children will offer the possibility of a better life potentially assured by one’s children in the future, and will allow one to maintain and expand one’s progeny.

Previous trends show that in many African countries young people are increasingly uninterested in contraceptive methods and the family planning process, with 9% of users aged 15-19 years between 1995 and 2020 in sub-Saharan Africa (15).

women’s empowerment the amount of wealth in a household had an influence on contraceptive use. When a woman resides in a household with a high wealth quantum, she becomes more likely to use a contraceptive than when she does not; this same trend is observed among women who have an occupation compared to those who have no occupation (16).

In the DRC, the number of living children a woman has seems to have a very slight influence on her ability to dispose of her income: the proportion of women who mainly decide on the use of their income increases from 27% among those with no living children to 28% among those with 1-2 children and then to 31% among those with 3-4 children. The difference by area of residence is greater: nearly five out of ten urban women (46%) decide how to use their income, compared to 21% in rural areas (17).

The desired number of children in a couple is also determined by the woman’s gynecological and obstetrical background. It is about pregnancies carried to term or not with the fetus having reached the age of viability or not. It can also be a personal experience or information on maternal-fetal mortality. The preference of the sex of the child is one of the factors that has a negative influence on contraception, some couples have more children simply in search of a female or male child, as the case may be, to meet a customary or cultural need. In Worawora Township, Ghana, 34% of couples with no children did not use contraception, while those with more than three children used a contraceptive method in 73% of cases (18).

Considering this background what are the factors that influence the use of modern contraceptives in Kinshasa, the capital of the Democratic Republic of Congo in 2020?

The objective of this study is to determine the factors associated with modern contraceptive use among women in Kinshasa, capital of Democratic Republic of Congo.

The specific objectives of this study are (i) Describe the sociodemographic factors of women in Kinshasa, (ii) Investigate factors related to the rate of using modern contraceptive, (iii) Assess the level of knowledge of family planning among non-menopausal adult women.

The research hypotheses are as follows: (i) Increasing age increases the use of modern contraceptives,

i. Involving the partner or husband in the family planning process increases modern contraceptive use,
ii. Increasing women’s knowledge of modern contraceptive methods increases their use, (iv) Low income levels lead to increased use of modern contraceptives, (v) The existence of a gynecological or obstetrical history increases modern contraceptive use.

## 2. MATERIALS AND METHOD

### 2.1 Study design

This is a cross-sectional study conducted in Kinshasa’s city, capital of the DRC, to determine the factors that influences modern contraceptives use in 2020, using secondary data coming from Performance Monitoring for Action (PMA) with support from Johns Hopkins Bloomberg School of public Health, Johns Hopkins Program for International Education in Gynecology and Obstetrics and Bill & Melinda Gates institute for population and reproductive health. With the collaboration of the Kinshasa high school of public health

### 2.2 Inclusion and exclusion criteria

All women of childbearing age, living in the study areas and aged between 15 and 49 years were included in this study. Excluded from this study were (i) Any women whose age was less than 15 years or more than 49 years, (ii) Any woman no longer living in the survey area or who was absent at the time of the survey and (iii) Any woman whose questionnaire is incomplete or who has not consented to be interviewed.

### 2.3 Sampling

A representative number of geographic clusters are sampled in collaboration with the National Institute of Statistics of the DR. Congo. 35 households were randomly selected using a multi-stage cluster design based on rural and urban strata after a complete listing of each enumeration area. The final sample included 2611 de facto women in Kinshasa aged 15-49 who completed the interviews. Data collection was conducted between December 2019 and February 2020

The following formula were used to determine the final sample size:

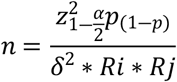

Where: n: sample size of women; Z: abscissa of normal curve (at α=0.05, Z=1.96); DEFF is the design effect (maximum: 3.0); P: estimated; δ: margin of error; Ri: individual response rate; Rh: household response rate.

### 2.4 Data analysis

The survey data was coded on an Excel file and analysis was done using Jamovi software version 2.2.5. Descriptive statistics and a chi-square test (P value < 0.05 for statistical significance.) were used to summarize and present the frequency distribution and percentages of variables. Logistic regression analysis with a 95% confidence interval and odds ratio value was used to estimate the association between the independent variables and the modern contraceptive use.

### 2.5 Ethical approval

During the surveys, the adolescents interviewed were considered emancipated minors and gave free and informed consent like all other survey participants. This approach received dual approval from the University of Kinshasa School of Public Health Ethics Committee under No. ESP/CE/030B/2019 of October 10, 2019, and the John Hopkins, Bloomberg School of Public Health Institutional Review Board Office of June 24, 2019 (IRB No. 00009677).

## 3. RESULTS

Analysis of the results on modern contraceptive use by women shows a steady trend in the choice of contraceptive method. Male condoms, implants, emergency contraception, injectable and pills are the most used methods by women (Table 1). Male condom was the most used method by respondents with 16.6% followed by implant with 15.4% of respondents. The emergency contraception use rate is 9.7% while injectable contraceptives are not widely used by respondents, only 6.6%, representing 72 women were users. The less used modern contraceptive methods are the pill, female sterilization and the IUD, with 3.6%, 1.3% and 0.3% of users respectively.

**Table 1.** Frequency distribution of use according to modern contraceptive type

| MC Method | Current user |  |
| --- | --- | --- |
|  | No | Yes |
| Emergency Contraceptive | 983(90.3 %) | 106(9.7 %) |
| Female sterilization | 1075(98.7 %) | 14(1.3 %) |
| Implant | 921(84.6 %) | 168(15.4 %) |
| Injectable | 1017(93.4 %) | 72(6.6 %) |
| Intra uterine device | 1085(99.6 %) | 4(0.4 %) |
| Male condoms | 908(83.4 %) | 181(16.6 %) |
| Pill | 1050(96.4 %) | 39(3.6 %) |

The prevalence of modern contraceptive use among women in Kinshasa during the survey period was 24.2%. The average age of participants was 28.3 years. The majority of participants were younger women aged 15-22 years representing 34.7%. Among them only 16.8% were users of modern contraceptive methods against 83.2% of non-users. Logistic regression analysis with 95% confidence interval, odds ratios and P:<0,05 for 9 predictors selected during this study show the following observation: Modern contraceptive use is strongly associated with age, partner involvement, household size, obstetric history, and the number of children desired (Table 2 and 3). Age significantly influences modern contraceptive use by women in Kinshasa during the study period.

**Table 2.** General characteristics of the study population

| Predictors | Current modern contraceptive user |  |  |
| --- | --- | --- | --- |
|  | n= 2611(%) | No (75.8) | Yes (24.2) |
| **Age Mean (SD)\*\*\*** | **28.3(9.47)** |  |  |
| 15-22 | 905(34.7 %) | 753(83.2 %) | 152(16.8 %) |
| 23-31 | 777(29.8 %) | 517(66.5 %) | 260(33.5 %) |
| 32-40 | 571(21.9 %) | 404(70.8 %) | 167(29.2 %) |
| 41-49 | 358(13.7 %) | 305(85.2 %) | 53(14.8 %) |
| **Marital status** |  |  |  |
| Unmarried | 1915(73.3 %) | 1466(76.6 %) | 449(23.4 %) |
| Married | 696(26.7 %) | 513(73.7 %) | 183(26.3 %) |
| **Woman's level education\*\*\*** |  |  |  |
| Never | 10(0.4 %) | 7(70.0 %) | 3(30.0 %) |
| Primary | 200(7.7 %) | 134(67.0 %) | 66(33.0 %) |
| Secondary | 1903(72.9 %) | 1479(77.7 %) | 424(22.3 %) |
| Tertiary | 498(19.1 %) | 359(72.1 %) | 139(27.9 %) |
| **Religion** |  |  |  |
| No religion | 86(3.3 %) | 60(69.8 %) | 26(30.2 %) |
| Catholic | 513(19.6 %) | 380(74.1 %) | 133(25.9 %) |
| Protestant | 285(10.9 %) | 206(72.3 %) | 79(27.7 %) |
| Kimbanguiste | 87(3.3 %) | 64(73.6 %) | 23(26.4 %) |
| Others christian | 1241(47.5 %) | 962(77.5 %) | 279(22.5 %) |
| Muslim | 44(1.7 %) | 34(77.3 %) | 10(22.7 %) |
| Others religion | 355(13.6 %) | 273(76.9 %) | 82(23.1 %) |
| Missing |  |  |  |
| **Partner involvement\*\*** | n= 1088 |  |  |
| No | 265(24.4 %) | 103(38.9 %) | 162(61.1 %) |
| Yes | 823(75.6 %) | 395(48.0 %) | 428(52.0 %) |
| **Occupation\*** |  |  |  |
| No | 1636(62.7 %) | 1263(77.2 %) | 373(22.8 %) |
| Yes | 974(37.3 %) | 715(73.4 %) | 259(26.6 %) |
| **Household members\*\*\*** |  |  |  |
| 1-3 | 1018(39.0 %) | 793(77.9 %) | 225(22.1 %) |
| 4-10 | 1352(51.8 %) | 1031(76.3 %) | 321(23.7 %) |
| 11-15 | 212(8.1 %) | 143(67.5 %) | 69(32.5 %)- |
| 16-20 | 29(1.1 %) | 12(41.4 %) | 17(58.6 %) |
| **Birth event\*\*\*** |  |  |  |
| No | 1065(40.8 %) | 898(84.3 %) | 167(15.7 %) |
| Yes | 1546(59.2 %) | 1081(69.9 %) | 465(30.1 %) |
| **Birth desire\*\*\*** |  |  |  |
| 0 | 1240(47.5 %) | 976(78.7 %) | 264(21.3 %) |
| 1-3 | 589(22.6 %) | 414(70.3 %) | 175(29.7 %) |
| 4-6 | 530(20.3 %) | 382(72.1 %) | 148(27.9 %) |
| 7 and over | 252(9.7 %) | 207(82.1 %) | 45(17.9 %) |

Note: \*P <0.05, \*\* P<0.01, \*\*\*P<0.001.

**Table 3.** Factors associated with modern contraceptive use among women in Kinshasa

| Predictors | Current modern contraceptive user |  |
| --- | --- | --- |
|  | n= 2368(%) | Odds (CI 95%) |
| **Age Mean (SD)** | 28.3(9.47) |  |
| 15-22 | 905(34.7 %) | 1 |
| 23-31 | 777(29.8 %) | 2.491(1.981-3.133)*** |
| 32-40 | 571(21.9 %) | 2.048(1.594-2.631)*** |
| 41-49 | 358(13.7 %) | 0.861(0.613-1.209) |
| **Marital status** |  |  |
| Unmarried | 1915(73.3 %) | 1 |
| Married | 696(26.7 %) | 1.165(0.954-1.421) |
| **Woman's level education** |  |  |
| Never | 10(0.4 %) | 1 |
| Primary | 200(7.7 %) | 1.149(0.288-4.59) |
| Secondary | 1903(72.9 %) | 0.669(0.172-2.6) |
| Tertiary | 498(19.1 %) | 0.903(0.23-3.54) |
| **Religion** |  |  |
| No religion | 86(3.3 %) | 1 |
| Catholic | 513(19.6 %) | 0.8080.49(1.333) |
| Protestant | 285(10.9 %) | 0.885(0.522-1.501) |
| Kimbanguiste | 87(3.3 %) | 0.829(0.428-1.608) |
| Others christian | 1241(47.5 %) | 0.669(0.415-1.08) |
| Muslim | 44(1.7 %) | 0.679(0.292-1.575) |
| Others religion | 355(13.6 %) | 0.693(0.411-1.168) |
| Missing | 41 |  |
| **Partner involvement** | n= 1088 |  |
| No | 265(24.4 %) | 1 |
| Yes | 823(75.6 %) | 0.689(0.519-0.914)* |
| **Occupation** | n=2610 |  |
| No | 1636(62.7 %) | 1 |
| Yes | 974(37.3 %) | 1.227(1.021-1.473)* |
| **Household members** |  |  |
| 1-3 | 1018(39.0 %) | 1 |
| 4-10 | 1352(51.8 %) | 1.097(0.904-1.332) |
| 11-15 | 212(8.1 %) | 1.701(1.231-2.349)** |
| 16-20 | 29(1.1 %) | 4.993(2.35-10.609)*** |
| **Birth event** |  |  |
| No | 1065(40.8 %) | 1 |
| Yes | 1546(59.2 %) | 2.313(1.898-2.819)*** |
| **Birth desire** |  |  |
| 0 | 1240(47.5 %) | 1.244(0.877-1.765) |
| 1-3 | 589(22.6 %) | 1.944(1.346-2.808)*** |
| 4-6 | 586(22.4 %) | 1.782(1.226-2.591)** |
| 7 and over | 196(7.5 %) | 1 |

Note: \*P <0.05, \*\* P<0.01, \*\*\*P<0.001.

Older women are statistically twice as likely to use modern contraceptives as women in the youngest age group, except for those aged 41-49, who showed a negative likelihood (0.86) of using modern contraceptives compared with their younger counterparts in the reference group, although this latter distribution is not statistically significant. Studies show that women’s likelihood of using modern contraceptives decreases as they age.

Married women were 1.16 times more likely to use modern contraceptives than unmarried women, even though this distribution is not statistically significant.

Having a primary education makes a woman 1,149 more likely to use modern contraceptives than a woman who has never studied. However, women with secondary education were less likely to use modern contraceptives than the reference women, while those with tertiary education had almost the same likelihood of use as women who had never studied.

Women with religion are less likely to use modern contraceptives than women with no religion. Catholic women have a negative odds (0.8) times of using modern contraceptives than women with no religion. The same is true for Protestant women (0.88), Muslim women (0.67) and women with other Christian religions (0.66). However, all these spiritual beliefs or religion did not show a statistically significant influence on modern contraceptive use.

Couple discussions, although very important, did not show a positive influence in the use of modern contraceptive methods. Women who discussed contraception with their partners or husbands were less likely to use modern contraceptives than those who did not discuss it with their partners (Odds:0.69).

Results from our Women’s Empowerment Study show that having an occupation increased the likelihood of using a contraceptive. women who had an occupation at the time of the study were 1,22 times more likely to use modern contraceptives than those with no occupation.

Household size significantly influences modern contraceptive use in Kinshasa. As household size increases, the likelihood of using modern contraceptives also increases. Women living in households with more than five people were more likely to use modern contraceptives than those living in households with fewer than six individuals. In other words, women in households of 16-20 people were 5 times more likely to use them than those in households of 1-5 people. Participants from households of 11-15 individuals were 1.7 times more likely to use modern contraceptives than those from reference group households.

Having given birth at least once positively and significantly influenced modern contraceptive use in study population. Women who had given birth at least once were 2,31 times more likely to use modern contraceptives than those who had never given birth. In the same vein, the desire for future birth influenced the choice of to use modern contraceptives. Women who wanted fewer children were more likely to use modern contraceptives than those who wanted more than 6 children. Women who no longer want children are 1.24 times more likely to use a modern contraceptive method than their counterparts who want 7 or more children. The same is true for those who want 1-3 children (1.94 times) and 4-6 children (1.78 times). All are more likely than women in the reference group.

Male condoms, implant, intrauterine device, emergency contraception, pills, injectable contraceptives, female sterilization, are the methods used by women in Kinshasa, in different proportions according to age. Distributions on emergency contraceptives, male condoms, implants and female sterilization are statically significant (Table 4)

**Tableau 4.** Frequency distribution of modern contraceptive methods by respondent age

| MC Method | Respondent's age |  |  |  |
| --- | --- | --- | --- | --- |
|  | 15-22 | 23-31 | 32-40 | 41-49 |
| EC *** | 38(35.8 %) | 48(45.3 %) | 18(17 %) | 2(1.9 %) |
| Female sterilization*** | 0(0%) | 0(0%) | 5(35.7 %) | 9 (64.3 %) |
| Implant*** | 21(12.5 %) | 60(35.7 %) | 67(39.9 %) | 20(11.9 %) |
| Injectable | 10(13.9 %) | 37(51.4 %) | 19(26.4 %) | 6(8.3 %) |
| Intra uterine device | 0(0%) | 1(25.0 %) | 3(75.0 %) | 0(0%) |
| Male condoms*** | 58(32%) | 76(42%) | 37(20.4 %) | 10(5.5 %) |
| Pill | 8(20.5 %) | 13(33.3 %) | 11(28.2 %) | 7(17.9 %) |

Note: \*P <0.05, \*\* P<0.01, \*\*\*P<0.001.

## 4. DISCUSSION

Contraceptive prevalence increased from 24.2%. These data show a certain evolution compared to the 6.5%, 5.4% and 8% recorded during the surveys conducted in 2003, 2010 and 2014 in the city of Minembwe, province of Sud-Kivu in DR Congo (19). This increase could be explained by the ease of access to family planning products and services, as well as the high population density in the city of Kinshasa compared to the Minembwe city. This improvement is still small compared to the number of non-users and the constant growth of the population, with a reproduction rate of 5.7 in DR Congo.

Age is a significant determinant of modern contraceptive use in Kinshasa. Analysis of the results shows that Older women are statistically twice as likely to use modern contraceptives as women in the youngest age group (15-22). These results are similar to those of Ahmad Kamran Osmani et al. who found that women under the age of 20 were significantly less likely to use contraceptives compared to their older counterparts in sub-Saharan Africa (8). This could be explained by the fact that younger women may be newly married and need children more than older women who may have already reached their desired number of children, hence the high use of modern contraceptives by women aged 23-40. In contrast, older women aged 41-49 showed a negative likelihood (0.86) of using modern contraceptives compared with their younger counterparts in the reference group. These results also show that this susceptibility decreases with increasing age; as age increases, women become less likely to use contraceptives. The occurrence of menopause in some cases and the likely preference for another, non-modern contraceptive method or the fact that some women still find the subject taboo could explain the low use by women aged 41-49.

The couple’s discussion of the appropriateness and value of using a contraceptive method to regulate births is a fundamental element to be taken into account in the study of influencing factors. During study period, discussion of contraception with the partner or husband had a negative influence on contraceptive use. Women who discussed contraception with their partners or husbands were less likely to use modern contraceptives than women who did not discussed with their partners or husbands (Odds: 0.689). This assertion could be explained by the fact that in African societies, particularly in the DR Congo, women, although submissive to men in a conjugal union, retain a certain autonomy on matters in which they are called upon to play a leading role in a context where many contraceptive methods have been designed for use by women. These results corroborate those of Freddy Rukema Kaniki, who points out that in 40% of cases husbands are responsible for the non-use of modern contraceptives by women in the province of South Kivu in DR Congo. (19)

Findings show a positive influence of increasing household size on women’s use of modern contraceptives. In households with more than 16 members, women are 5 times more likely to use modern contraceptives than women in households with 1-5 members. As household size increases, women are more likely to use modern contraceptives. These results are consistent with those of a study of household structure and contraceptive use in Nigeria, which found that in households with more than five members, women were 1.55 times more likely to use contraceptives than in households with fewer than five members (20). African societies are marked by the hospitality of wealthy family members towards other members who lack the means to survive. It is in this context that we find in several households sometimes several members other than the couple’s own children. The social and economic conditions of the population in general and of households in particular force couples, especially women, to think more carefully before considering a new pregnancy.

There is a significant association between a woman’s obstetrical history, her desire for future birth, and the use of a modern contraceptive method.

Women with at least one obstetrical history (birth) were 2.31 times more likely to use modern contraceptives than women who never given birth. In Zambia Lasong J et al. also found that women who had 5 or more deliveries were 8.02 times more likely than women who had never delivered (21). This result could be explained by the fact that according to many social cultures in DR Congo, it is not advisable to have a child or to be sexually active before marriage. Considering that in this study most of women are not married, they would probably refuse to become pregnant again after an obstetrical history.

Women who want fewer future births used modern contraceptives more than women who want more than 6 children. women who did not want any more children were 1.24 times more likely to use modern contraceptives than those who wanted 7 and more, while women who want 1-3 children are twice as likely to use as those who want 7 or more (Odds: 1.944). These results are similar to those of Ahmed Islam who found in a study conducted in Bangladesh that women who did not want any more children were 2,504 times more likely to use modern contraceptives than those who wanted more. The explanation for this similarity is that the need for future births is matched by the financial capacity of couples to ensure the growth and development of children, which is not the case in many poor countries such as DR Congo and Bangladesh. (22)

## Strengths and limitations

Strengths of this study include a cross-sectional study design with a relatively large sample size (n=2611) providing information on recent data on most determinants of modern contraceptive use in Kinshasa, with probable integrity on key points, thus allowing access to several results and exhibitions.

Female age, occupation, and obstetrical history may play a role in the prevalence of modern contraception.

Various limitations should be taken into account when interpreting the results of study:

i. This study allows for the assessment of the association between the predictors and the dependent variable, but could not allow for the assessment of the causal relationship.
ii. These results may not be generalized over the whole city of Kinshasa with its 15 million inhabitants considering ethnic and cultural diversity.
iii. Not enough information about the culture of and residences of respondents.

However, this study could provide a basis for future research opportunities.

## 5. CONCLUSION

Age is a significant determinant of modern contraceptive use. Older women are more likely to use modern contraception than women in the reference group except the oldest, and their likelihood of use diminishes as they age. Women are most likely to use modern contraceptives when they do not discuss the topic with their husbands or partners. As household size increases, women’s use of modern contraceptives positively is affected. The same is applicable for women who have given birth at least compared to those who have never given birth. When a woman wants fewer children, she is more likely to use these contraceptives than when she wants more. Male condoms, emergency contraceptives and pills were used more by younger women while the most common contraceptive method used only by older women is female sterilization.

Political Implication: In a country with a rapidly growing population such as the DRC with higher reproduction rate, health policies on family planning should in the future focus on raising awareness among young women. The government should introduce new laws and strengthen existing laws on outreach and school-based counseling for girls by expanding the scope and capacity of the national reproductive health program and the school health program. This policy should be built on the use of male condoms, emergency contraception and pills by the girl as the main means of contraception.

Furthermore, accelerate the development, implementation or strengthening of a program to promote the sexual and reproductive health for women of reproductive age in order to achieve the sustainable development goals, including those related to sexual and reproductive health by 2030.

## Data Availability

The data used in this research are those of PMA (Programme monitoring for action) after a proper data request procedure. Below is the link that allowed us to download the database in excel format. https://datalab.pmadata.org/dataset/doi%3A1034976x3bk-0w75 https://datalab.pmadata.org/dashboard

## ACKNOWLEDGMENTS

My deepest thanks go:

to God Almighty who made this moment possible,

to Yonsei University and all the administrative and academic team members,

to my father MOKEKE MOLUA Alphonse and to my late mother AKOLA MIKOLANGO Venance who rests in eternal bliss, as well as to my other family members,

to my dear beloved wife NTUMBA KIALA Sarah, a dynamic woman, for her love and encouragement, to all my friends and acquaintances for their support

---

**Figure.**
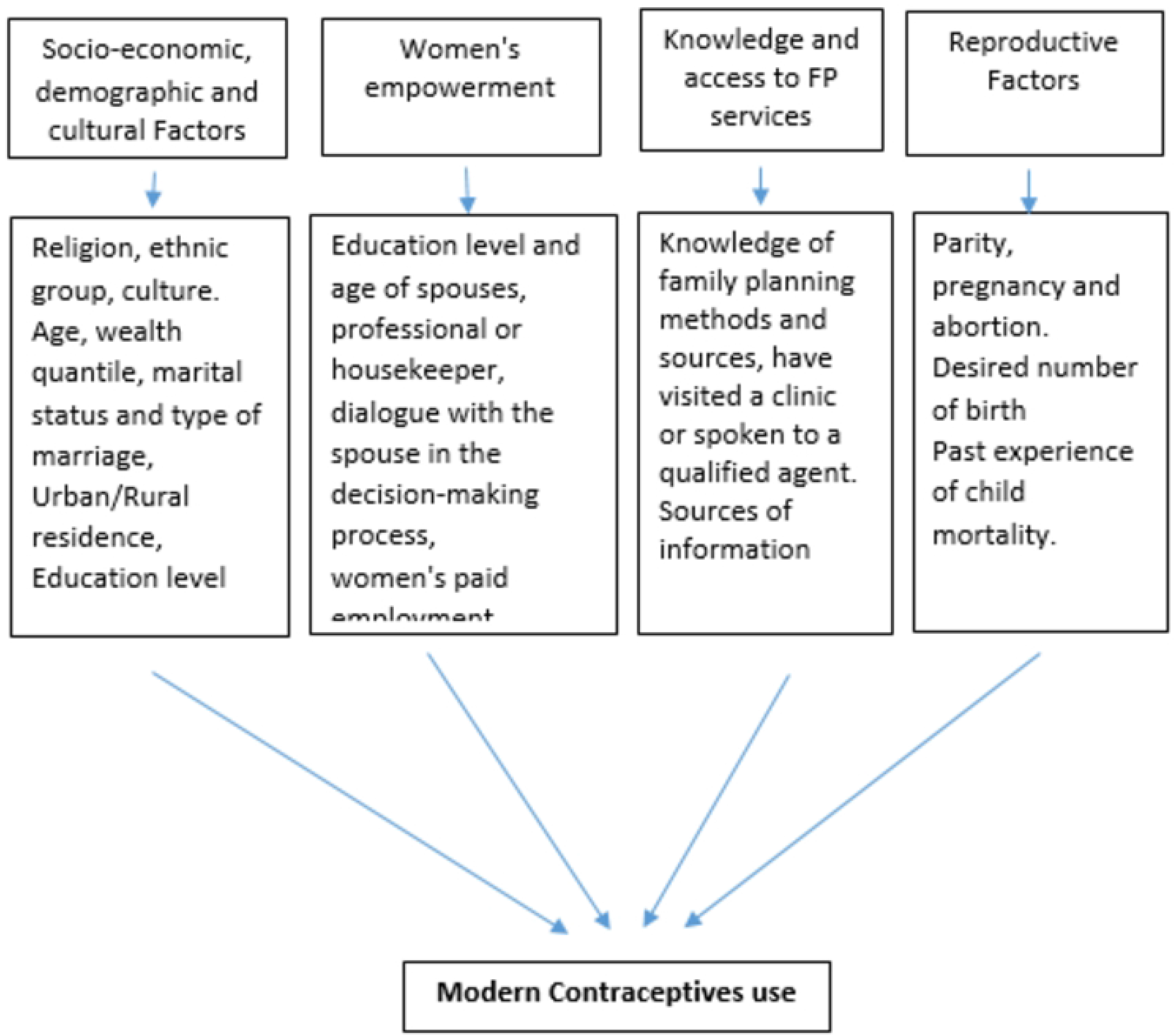

## REFERENCES

1. Shaw D. The ABCs of family planning Canada: THE GLOBE AND MAIL; 2010 [cited 2022. Available from: https://www.theglobeandmail.com/opinion/the-abcs-of-family-planning/article4310750/.

2. Gateway B. BIBLE English Standard Version (ESV): Bible Gateway; 2022.

3. Ethiopia FDRo. HEAT Health Education and Training HEAT in Africa Federal Democratic Republic of Ethiopia Ministry of Health Family Planning Blended Learning Module for the Health Extension Programme. In: Health Mo, editor. http://academia.edu: Sileshi A Garoma; 2010. p. 148.

4. Blogger G. Lindsey Horvath, community activist in Los Angeles and a Global Coordinator for the V-Day One Billion Rising Campaign United Nations Foundation: United Nations Foundation; 2014 [Available from: https://unfoundation.org/blog/post/end-all-injustice-against-women/.

5. Geneva WHO, 2011. Unsafe abortion incidence and mortality: global and regional estimates of incidence of unsafe abortion and associated mortality in 2008. World Health organization, Information sheet: World Health organization; 2008.

6. Osmani AK, Reyer JA, Osmani AR, Hamajima N. Factors influencing contraceptive use among women in Afghanistan: secondary analysis of Afghanistan health survey 2012. Nagoya J Med Sci. 2015;77.

7. La contraception: la clé pour atteindre les ODD 2030 [press release]. FIGO web: FIGO, Londres, 20 septembre 2018.

8. Ahmed S, Choi Y, Rimon JG, Alzouma S, Gichangi P, Guiella G, et al. Trends in contraceptive prevalence rates in sub-Saharan Africa since the 2012 London Summit on Family Planning: results from repeated cross-sectional surveys. Lancet Glob Health. 2019;7(7):e904–e11.

9. The World Bank group W. Mortality rate, infant, male (per 1,000 live births) -Congo, Dem. Rep. 2022 [Available from: https://data.worldbank.org/indicator/SP.DYN.IMRT.MA.IN?locations=CD.

10. 2022 WPRKP. World Population Review. Kinshasa Population 2022. Retrieved Nov 27, 2022, from http://www.worldpopulationreview.com xRetrieved Nov 27, 2022.

11. Augustin Kadiata Bukasa EKMac. Daily Life of Adolescent Girls who Experienced Early Motherhood in the City of Kinshasa. Scholars International Journal of Obstetrics and Gynecology (SIJOG). 2022.

12. Family planning F. Congo, FP2020 Core Indicator Summary Sheet: 2019-2020 Annual Progress Report. 2020.

13. Ministry of health D. Plan Stratégique pour la planification familiale 2021-2025. 2021-2025.

14. Ministère de la Santé Publique -MSP/RDC. Loi n° 18/035 du 13 décembre 2018 fixant les principes fondamentaux relatifs à l’organisation de la Santé publique. Journal Officiel de la RDC: JOS 31.12.2018; 2018. p. 23.

15. Boadu I. Coverage and determinants of modern contraceptive use in sub-Saharan Africa: further analysis of demographic and health surveys. Reproductive Health. 2022;19(1):18.

16. Yaya S, Uthman OA, Ekholuenetale M, Bishwajit G. Women empowerment as an enabling factor of contraceptive use in sub-Saharan Africa: a multilevel analysis of cross-sectional surveys of 32 countries. Reproductive Health. 2018;15(1):214.

17. Ministère du Plan et Suivi de la Mise en œuvre de la Révolution de la Modernité -MPSMRM/Congo, Ministère de la Santé Publique -MSP/Congo, ICF International. République Démocratique du Congo Enquête Démographique et de Santé (EDS-RDC) 2013-2014. Rockville, Maryland, USA: MPSMRM, MSP, and ICF International; 2014.

18. Manortey S, Lotsu P. Factors affecting contraceptive use among reproductive aged women: a case study in Worawora township, ghana. J Sci Res Rep. 2017.

19. Kaniki FR. Factors influencing the use of modern contraceptive methods among rural women of child bearing age in the Democratic Republic of the Congo. J Family Med Prim Care. 2019;8(8):2582–6.

20. Fadeyibi O, Alade M, Adebayo S, Erinfolami T, Mustapha F, Yaradua S. Household Structure and Contraceptive Use in Nigeria. Frontiers in Global Women’s Health. 2022;3.

21. Lasong J, Zhang Y, Gebremedhin SA, Opoku S, Abaidoo CS, Mkandawire T, et al. Determinants of modern contraceptive use among married women of reproductive age: a cross-sectional study in rural Zambia. BMJ Open. 2020;10(3):e030980.

22. Islam AZ, Mondal MNI, Khatun ML, Rahman MM, Islam MR, Mostofa MG, et al. Prevalence and determinants of contraceptive use among employed and unemployed women in Bangladesh. Int J MCH AIDS. 2016;5.

